# The critical role of infection prevention overlooked in Ethiopia, only one-half of health-care workers had safe practice: A Systematic Review and Meta-Analysis

**DOI:** 10.1101/2020.05.07.20094672

**Authors:** Biniyam Sahiledengle, Yohannes Tekalegn, Demelash Woldeyohannes

**Affiliations:** Department of Public Health, Madda Walabu University Goba Referral Hospital, Bale Goba, Ethiopia

**Keywords:** Infection prevention, standard precaution, healthcare-associated infections, healthcare workers, Ethiopia, systematic review, meta-analysis

## Abstract

**Background:** Effective infection prevention and control measures, such as such hand hygiene, the use of personal protective equipment, instrument processing, safe injection, and safe disposal of infectious wastes in the healthcare facilities maximize patient outcomes and are essential to providing effective, efficient, and quality health care services. In Ethiopia, findings regarding infection prevention practices among healthcare workers have been highly variable and uncertain. Therefore, this systematic review and meta-analysis estimate the pooled prevalence of safe infection prevention practices and summarize the associated factors among healthcare workers in Ethiopia.

**Methods:** PubMed, Science Direct, Google Scholar, and the Cochrane library were systematically searched. We included all observational studies reporting the prevalence of safe infection prevention practices among healthcare workers in Ethiopia. Two authors independently extracted all necessary data using a standardized data extraction format. Qualitative and quantitative analyses were employed. The Cochrane Q test statistics and I^2^ tests were used to assess the heterogeneity of the studies. A random-effects meta-analysis model was used to estimate the pooled prevalence of safe infection prevention practice.

**Results:** Of the 187 articles identified through our search, 10 studies fulfilled the inclusion criteria and were included in the meta-analysis. The pooled prevalence of safe infection prevention practice in Ethiopia was 52.2% (95%CI: 40.9-63.4). The highest prevalence of safe practice was observed in Addis Ababa (capital city) 66.2% (95%CI: 60.6-71.8), followed by Amhara region 54.6% (95%CI: 51.1-58.1), and then Oromia region 48.5% (95%CI: 24.2-72.8), and the least safe practices were reported from South Nation Nationalities and People (SNNP) and Tigray regions with a pooled prevalence of 39.4% (95%CI: 13.9-64.8). In our qualitative syntheses, healthcare workers socio-demographic factors (young age, female gender), behavioral-related factors (being knowledgeable and having a positive attitude towards infection prevention), and healthcare facility-related factors (presence of running water supply, availability of infection prevention guideline, and receiving training) were important variables associated with safe infection prevention practice.

**Conclusions:** Only half of the healthcare workers in Ethiopia practiced safe infection prevention. Furthermore, the study found out that there were regional and professional variations in the prevalence of safe infection prevention practices. Therefore, the need to step-up efforts to intensify the current national infection prevention and patient safety initiative as key policy direction is 41 strongly recommended, along with more attempts to increase healthcare worker’s adherence towards infection prevention guidelines.

## Background

Effective infection prevention and control measures, such as proper hand hygiene, the use of personal protective equipment (PPE), environmental cleaning, instrument processing, safe injection, and safe disposal of infectious wastes in the healthcare facility maximize patient outcomes and are essential to providing effective, efficient, and quality health care services. [1-3].

Worldwide, HAIs affecting the quality of care of hundreds of millions of patients every year, contributing to increased morbidity, mortality, and substantial healthcare cost [1,2]. According to the World Health Organization (WHO), at any point in time in every hundred hospitalized patients, ten in developing countries and seven in developed countries will acquire at least one HAI [3]. Data from the European Centre for Disease Prevention and Control (ECDC) surveillance reports showed that the prevalence of patients with at least one HAI in acute care hospitals in Europe was 6.0% (country range 2.3%–10.8%)[4]. The Centre for Disease Prevention and Control (CDC) estimates 2 million patients who will suffer from HAIs every year in the United States (US), and a near hundred thousand of them die [5], which cost as much as 4.25 billion United States dollars [6].

In developing countries, the burden of HAIs is largely unknown, and in most cases, is underestimated due to the lack of proper surveillance. Studies conducted in low-income settings showed that the prevalence of HAIs varies from 5.7% to 19.1%, with a pooled prevalence of 10.1% [7]; and the cumulative incidence range is from 5.7% to 45.8% [8]. Remarkably, in many sub-Saharan African countries including Ethiopia the true prevalence of HAIs is still uncertain [8], and adherence towards infection prevention recommendations among healthcare workers (HCWs) is poor [9-12]. Known in this region of Africa, the risk of occupational injury and exposure to blood-borne infections is on the rise. A review by Auta et al showed that the estimated pooled lifetime prevalence on occupational exposure to body fluids among HCWs in 21 countries in Africa was 65.7% [13]. The only sustainable, practical, and long-term solution to reduce the problems of HAIs, and occupational exposures lie on actions to implement safe infection prevention practices in all healthcare facilities [3,9,10].

Ethiopia, similar to other African countries, does not have a well-described report on the burden of HAIs. Few HAI studies are available that focus on the incidents and prevalence of HAIs [14-17]. A study by Endalafer et al reported the overall incidence of nosocomial infections in a tertiary hospital in Ethiopia was 35.8/100 patients, which is enormous [14]. Moreover, a high prevalence of HAIs has reported from all corners of the country, 15.4% in north Ethiopia [15], 11.4%-19.4% in southwest Ethiopia [16,17], and 16.4% in central Ethiopia [18]. The evidence available further suggests that inadequate infection prevention practices among HCWs are frequent [19, 20].

Sufficient evidence that demonstrated the role of infection prevention on the reduction of HAIs [21-23]- for example, Sickbert et al, in their study reported was an improvement in hand hygiene compliance by 10%, which associated a significant reduction in overall HAIs [22]. It is also reported that effective infection prevention and control reduce HAIs by at least 30% [21]. Despite these facts-in many low-income settings, lack of well-trained HCWs, lack of infection prevention and control policies, and technical guidelines made the problem even worse [9,15,24-28].

To maximize the prevention of HAIs at the national level in Ethiopia, there has been a growing recognition of the need for guidance on the documents, and in 2012, the publication of the second national infection prevention and patient safety guidelines was released. From that day on, considerable progress has been made in understanding the basic principles, acceptance, and use of evidence-based infection prevention practices in Ethiopia [9]. Despite the effort underway, studies reported inconsistent findings regarding HCWs infection prevention practice in Ethiopia [19,20,27,29-33]. Although the reporting of such practices is important for the prevention and control of HAIs and improving quality of care, the previously conducted studies had many differences in the geographical regions and preceded remarkable variations in the reported practices. Due to the aforementioned reason, we conducted a systematic review and meta-analysis of observational studies to estimate the pooled prevalence of safe infection prevention practices among HCWs in Ethiopia. Also, we aim to summarize descriptively the factors that were associated with safe practice.

## Materials and methods

### Search strategy

This systematic review and meta-analysis analyzed the pooled prevalence of safe infection prevention practices among HCWs in Ethiopia. In addition, we qualitatively analyzed the factors that were associated with safe infection prevention practices from the included studies based on the articles published in national and international journals. The protocol for this review was registered in the International Prospective Register of Systematic Reviews (PROSPERO), the University of York Centre for Reviews and Dissemination (record ID: CRD42019129167, on the 31^st^ May 2019).

Databases including PubMed/MEDLINE, Science Direct, Cochrane Library, and Google Scholar were systematically searched. Also, we screened at the references lists of identified articles to detect and identify additional relevant studies for this review. Furthermore, to find unpublished papers relevant to this systematic review and meta-analysis, Addis Ababa University Digital Library were searched. The search for the literatures was conducted between the 15^th^ of April to the 31^st^ of May, 2019. The following terms and keywords were applied for PubMed/MEDLINE search: (infection prevention OR infection control OR standard precaution OR practice) AND (healthcare workers OR health workers OR health personnel OR healthcare providers) AND (health facilities OR hospitals OR public health facilities) AND (Ethiopia) as well as all possible combinations of these terms. For the other electronic databases, we used database-specific subject headings linked with the above terms and keywords used in PubMed. This review is reported according to the Preferred Reporting Items for Systematic Reviews and Meta-Analyses (PRISMA) guidelines [34] (**S1 File)**. The search strategy is provided in **S2 File and S3 File**.

**S1 File. Table. PRISMA Checklist**

**S2 File. Search Strategy (Full searching strategies for PubMed)**

**S3 File. Search Strategy (Example Google scholar)**

### Inclusion criteria

- Study design: observational studies
- Population: only studies involving healthcare workers
- Language: articles published in the English language
- Reported condition: studies that reported the overall healthcare workers infection prevention practice
- Availability of full texts
- Study area: studies conducted in Ethiopia

### Exclusion criteria

Articles with the following characteristics were excluded from this review

- Studies whose full data were not accessible even after requests from the authors
- Studies which did not report the overall prevalence of infection prevention practices
- Studies conducted on medical, interns, health science students and housekeeping staffs
- Qualitative studies, reviews, commentaries, editorials, letters, interventional studies, and other opinion papers
- Excluded published articles with unclear methods

### The outcome of the study

These systematic reviews have two main outcomes. Prevalence of safe infection prevention practices in Ethiopia, as the primary outcome variable of this study, is defined as the overall correct practice of the core components of infection prevention that include hand hygiene, utilization of personal protective equipment, reusable medical equipment processing, healthcare waste management, tuberculosis infection control, and safe injection and medication practices. The prevalence was computed by dividing the number of healthcare workers reported correct/acceptable infection prevention practices by the total number of healthcare workers (sample size), and multiplied by 100. The second outcome of this study was to summarize descriptively the factors that were associated with safe infection prevention practices in Ethiopia from the included studies.

### Data extraction

Two investigators (BS and YT) independently extracted the data from the studies included in our analysis as recommended by PRISMA guidelines [34]. The data were extracted using a standard data extraction forms. The following information were extracted from the selected studies: first author’s name, year of publication, the type of study design, study setting, study population, sample size, sampling methods, the magnitude of infection prevention practice, infection prevention components assessed, response rate, and region.

### Quality assessment

The assessment of methodological quality was carried out independently by two reviewers using the Newcastle-Ottawa scale (NOS) [35]. This scale has three sections: 1^st^ selection (maximum 5 stars), (2) comparability between groups (maximum 2 stars), and (3) outcome assessment (maximum 3 stars). In summary, the maximum possible score was 10 stars, which represented the highest methodological quality. The two authors (BS and YT) independently assessed the quality of each original study using the quality assessment tool. Any disagreements during the data extraction were resolved through discussion and consensus. Finally, any article with a scale of greater than or equal to ≥ 7 out of 10 was included in this Systematic Review and Meta-analysis. A detailed scoring result was described in the supplementary file (**S4 File**).

**S14 File. Table: Methodology quality assessment of included and excluded studies**

### Data analysis and synthesis

Data extractions were done using Microsoft Excel spreadsheet, then analyses were done using STATA version 14 statistical software. The descriptive data were presented using a table to describe the characteristics of each primary study. The standard errors for each original study were calculated using the binomial distribution formula. The presence of heterogeneity among the reported prevalence was assessed by computing p-values for the Cochran Q test and I^2^ test. Cochran’s Q test was used to test the null hypothesis of no significant heterogeneity across the studies [36]. Although there can be no absolute rule for when heterogeneity becomes important, Higgins et al. tentatively suggested low for I^2^ values between 25%–50%, moderate for 50%–75%, and high for ≥75% [36]. Subgroup analysis was done by the region where primary studies were conducted, publication year, sample size, sampling method, and type of healthcare facility.

Publication bias was assessed using a funnel plot. In the absence of publication bias, the plot resembles a symmetrical large inverted funnel. Egger's weighted regression and Begg’s rank correlation tests were used in checking the publication bias (P < 0.05), considered statistically significant [37]. We also conducted a leave-one-out sensitivity analysis to appraise the main studies that exerted an important impact on between-study heterogeneity.

## Results

### Identification of studies

For this review, one hundred and eighty-seven studies were identified in the initial search. Of these, 118 were excluded during the evaluation of the title and abstract. And based on the inclusion and exclusion criteria, a total of 10 studies were included in the final systematic review and meta-analysis (Fig 1).

**Fig 1:**
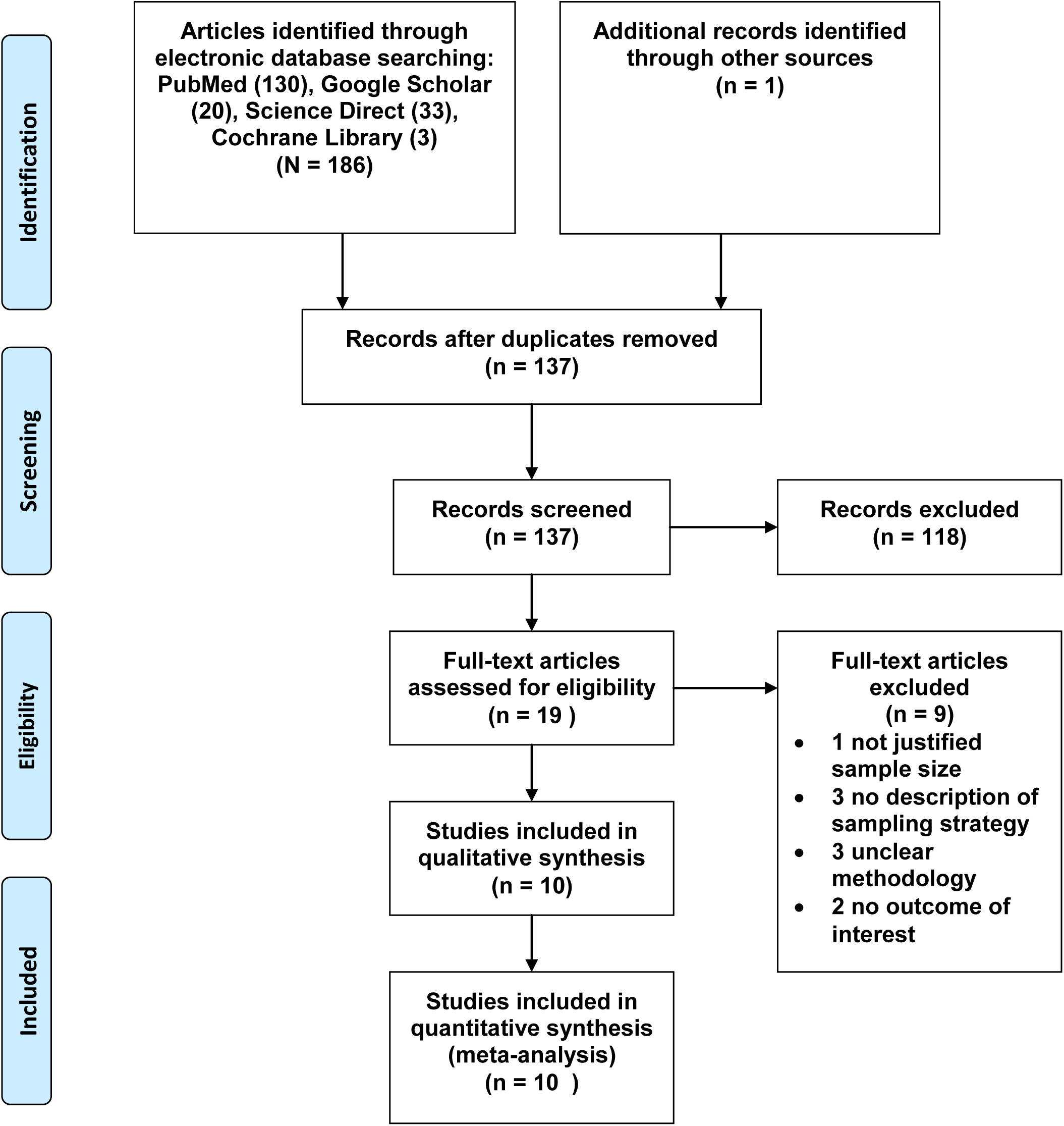
PRISMA flow chart of review search.

### Characteristics of included studies

A total of 10 articles [19,20,27,31,33,38-42] were included in meta-analysis. The study sample included 3,510 samples with a mean of 351 samples in each study. The maximum and minimum sample sizes were found in studies conducted by Geberemariyam BS., et al (648 samples) in the Oromia region [20] and Abreha N., et al. (108 samples) in Addis Ababa (the capital city of Ethiopia) [41]. Selected studies were conducted between 2014 and 2019. All the included studies were cross-sectional by design. With regards to regional distribution, about (30%) of the studies were conducted in Addis Ababa [27,40,41]. The prevalence of safe infection prevention practices ranged between 15% [33], and 72.5% [41] in South Nation Nationalities and People (SNNPs) Region and Addis Ababa, respectively. Concerning the quality score, all included studies were of a reputable methodological quality score of 7 from a total of 10-point (**Table 1)**.

**Table 1:**
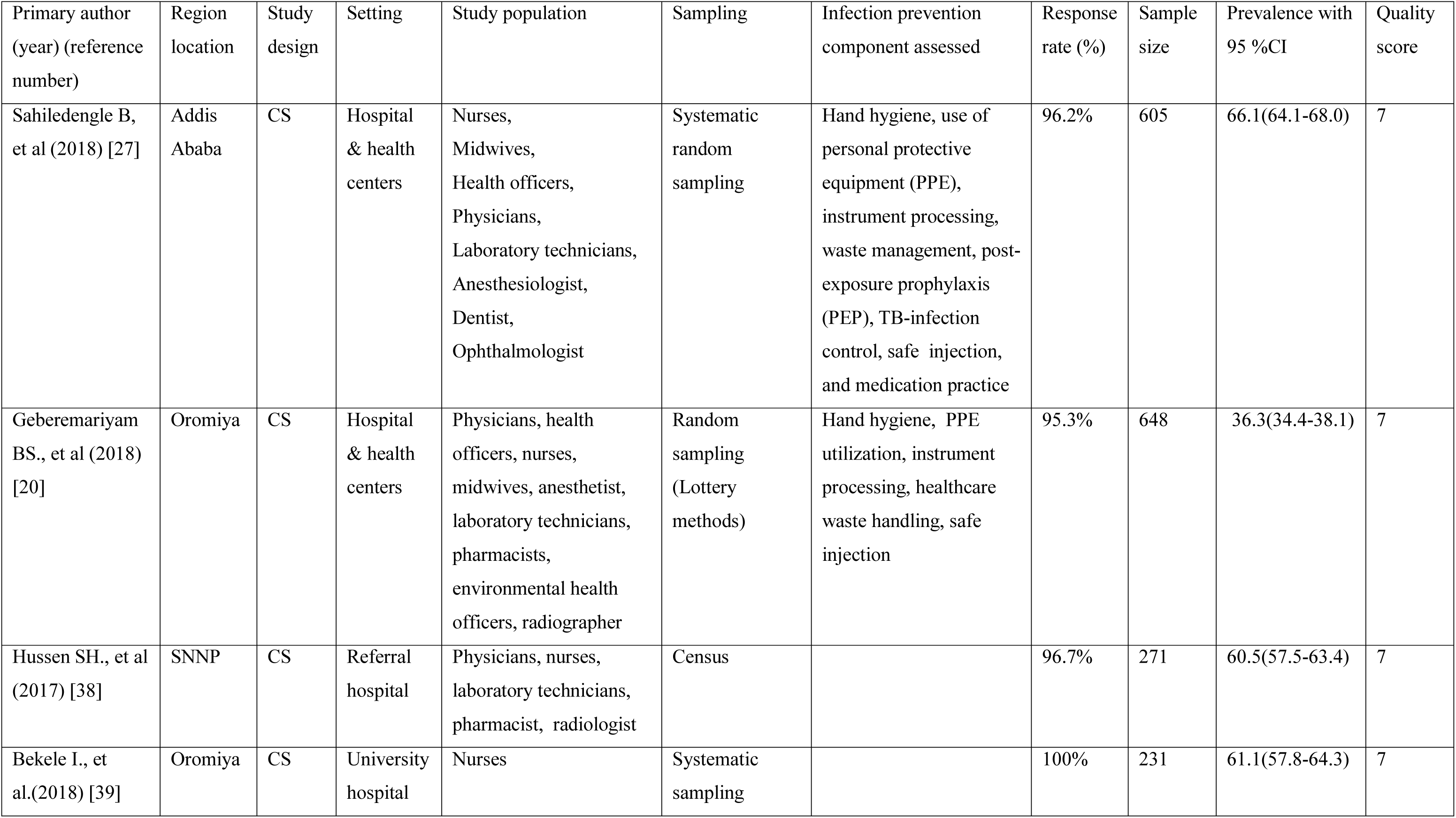
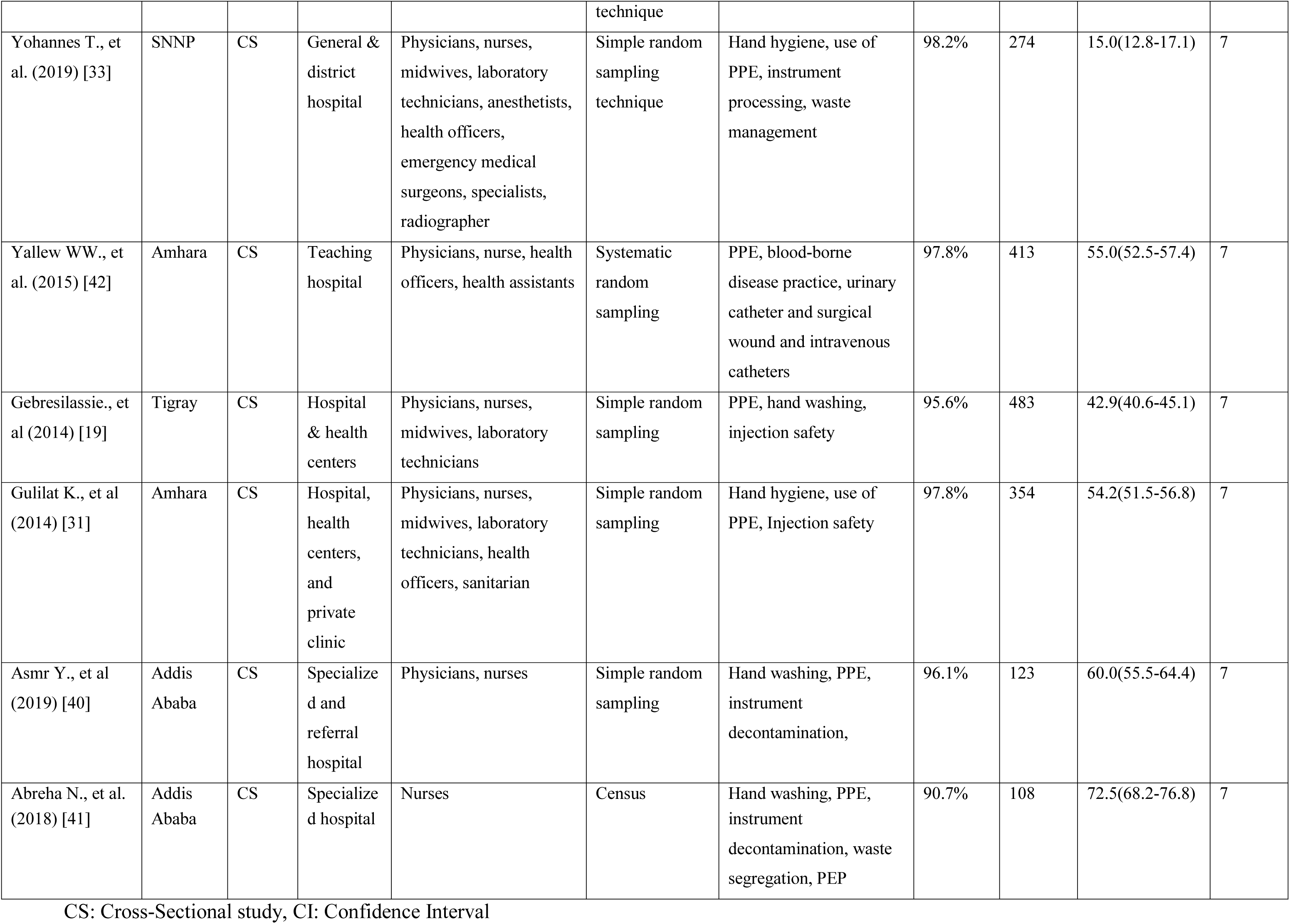
List of studies included that shows the prevalence of safe infection prevention practice among healthcare workers in Ethiopian, a systematic review and meta-analysis, Ethiopia, 2014-2019.

## Meta-Analysis

### Prevalence of safe infection prevention practices

A total of ten studies were included in the meta-analysis. From these studies, the pooled prevalence of safe infection prevention practices in Ethiopia was 52.2% (95%CI: 40.9-63.4). A significant higher heterogeneity among the ten included studies was found (I^2^=98.0%; Q=453.55, Variance Tau-squared = 319.63, p<0.001). Due to the existence of significant heterogeneity, we used a random-effect meta-analysis model to estimate pooled prevalence (**Fig 2**). According to the sensitivity analysis, there was no single influential study that significantly accounted for it (**Table 2)**.

**Fig 2:**
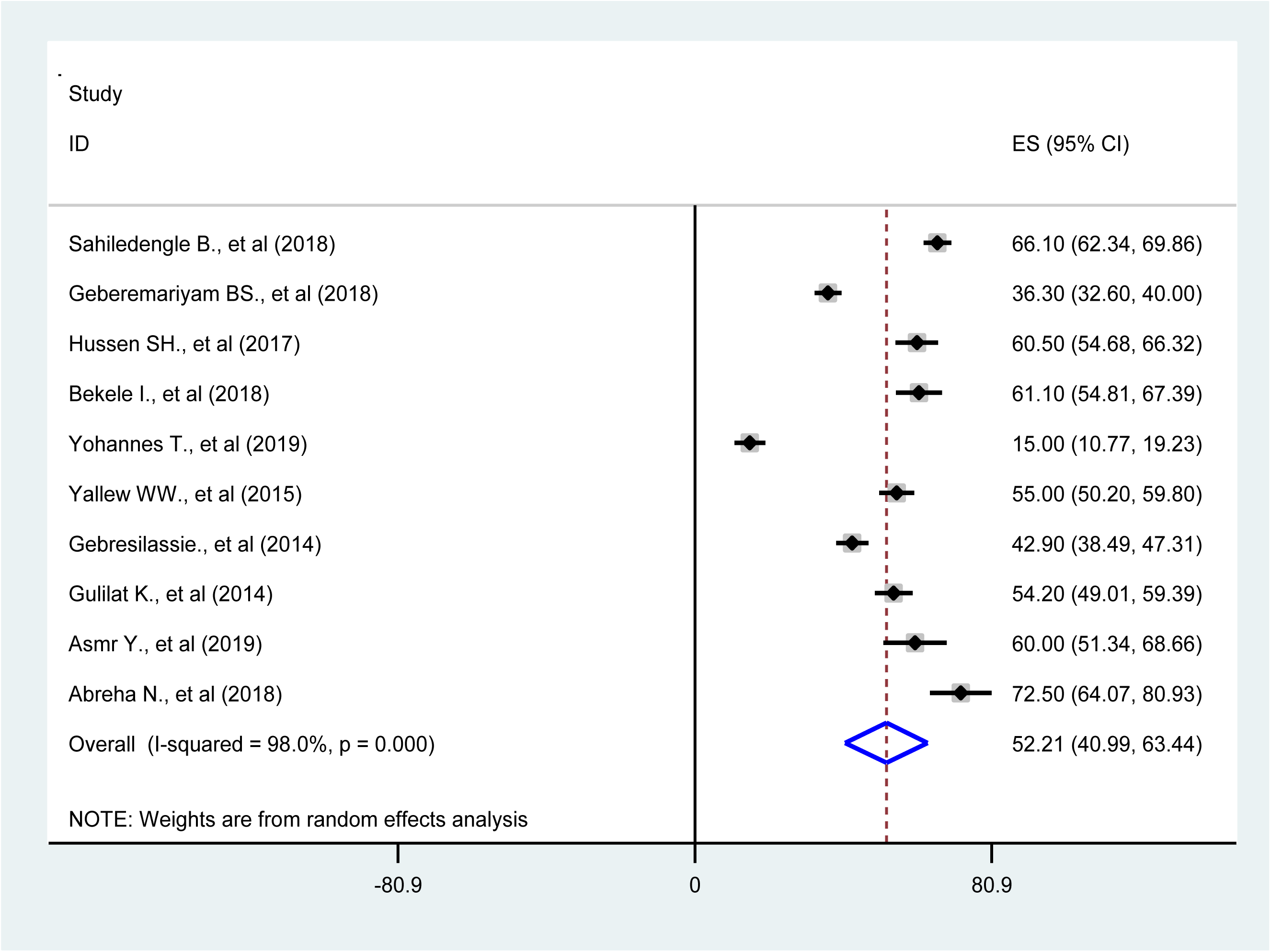
Forest plot of the pooled prevalence of safe infection prevention practice in Ethiopia, 2014-2019

**Table 2.** Sensitivity analysis of prevalence for each study being removed at a time: Prevalence 227 and 95% confidence interval of infection prevention practice in Ethiopia, 2014-2019.

| Study excluded | Prevalence | 95% CI | $I^2$ (%) | Q | p-value |
| --- | --- | --- | --- | --- | --- |
| Sahiledengle B., et al (2018) [27] | 50.6 | 39.1-62.1 | 97.7 | 348.0 | $p<0.001$ |
| Geberemariam BS., et al (2018) [20] | 54.0 | 41.6-66.4 | 98.0 | 405.8 | $p<0.001$ |
| Hussen SH., et al (2017) [38] | 51.3 | 39.1-63.4 | 98.2 | 434.9 | $p<0.001$ |
| Bekele I., et al (2018) [39] | 51.2 | 39.1-63.3 | 98.2 | 436.2 | $p<0.001$ |
| Yohannes T., et al (2019) [33] | 56.2 | 48.1-64.4 | 95.6 | 181.3 | $p<0.001$ |
| Yallew WW., et al (2015) [42] | 51.9 | 39.4-64.4 | 98.2 | 444.8 | $p<0.001$ |
| Gebresilassie., et al (2014) [19] | 53.2 | 40.5-65.9 | 98.2 | 447.3 | $p<0.001$ |
| Gulilat K., et al (2014) [31] | 52.0 | 39.5-64.4 | 98.2 | 447.8 | $p<0.001$ |
| Asmr Y., et al (2019) [40] | 51.3 | 39.4-63.3 | 98.2 | 446.1 | $p<0.001$ |
| Abreha N., et al (2018) [41] | 50.0 | 38.3-61.6 | 98.1 | 420.3 | $p<0.001$ |

## Subgroup Analyses

### The subgroup analyses of infection prevention practice prevalence

The results of the subgroup analysis showed that the pooled prevalence of safe infection prevention practices were highest in Addis Ababa (capital city) 66.2% (95%CI: 60.6-71.8) [I^2^= 51.4%, p=0.128], and 54.6% (95%CI: 51.1-58.1) [I^2^= 0.0%, p=0.825] in Amhara Region; 48.5% (95%CI: 24.2-72.8) in Oromia Regional State; and the least safe practices were reported from other regions (SNNP and Tigray regions) with pooled prevalence of 39.4% (95%CI: 13.9-64.8). A considerable heterogeneity was also found [I^2^= 97.7%; p<0.001]; and [I^2^= 98.8%; p<0.001] for the Oromia Regional State, and other regions (SNNP and Tigray), respectively. The prevalence of infection prevention practices was analyzed separately for either nurses or all other healthcare workers. The present findings show the prevalence of safe infection prevention practices more in studies conducted exclusively on nurses than in other health care workers (66.4% vs. 48.6%). We also conducted a subgroup analysis based on the study setting. The pooled prevalence of safe infection prevention practice showed more in studies conducted exclusively in hospitals than in those that include health centers (53.5% vs. 49.8%). More details on the prevalence of safe infection prevention practices for subgroups are presented in **Table 3**.

**Table 3:**
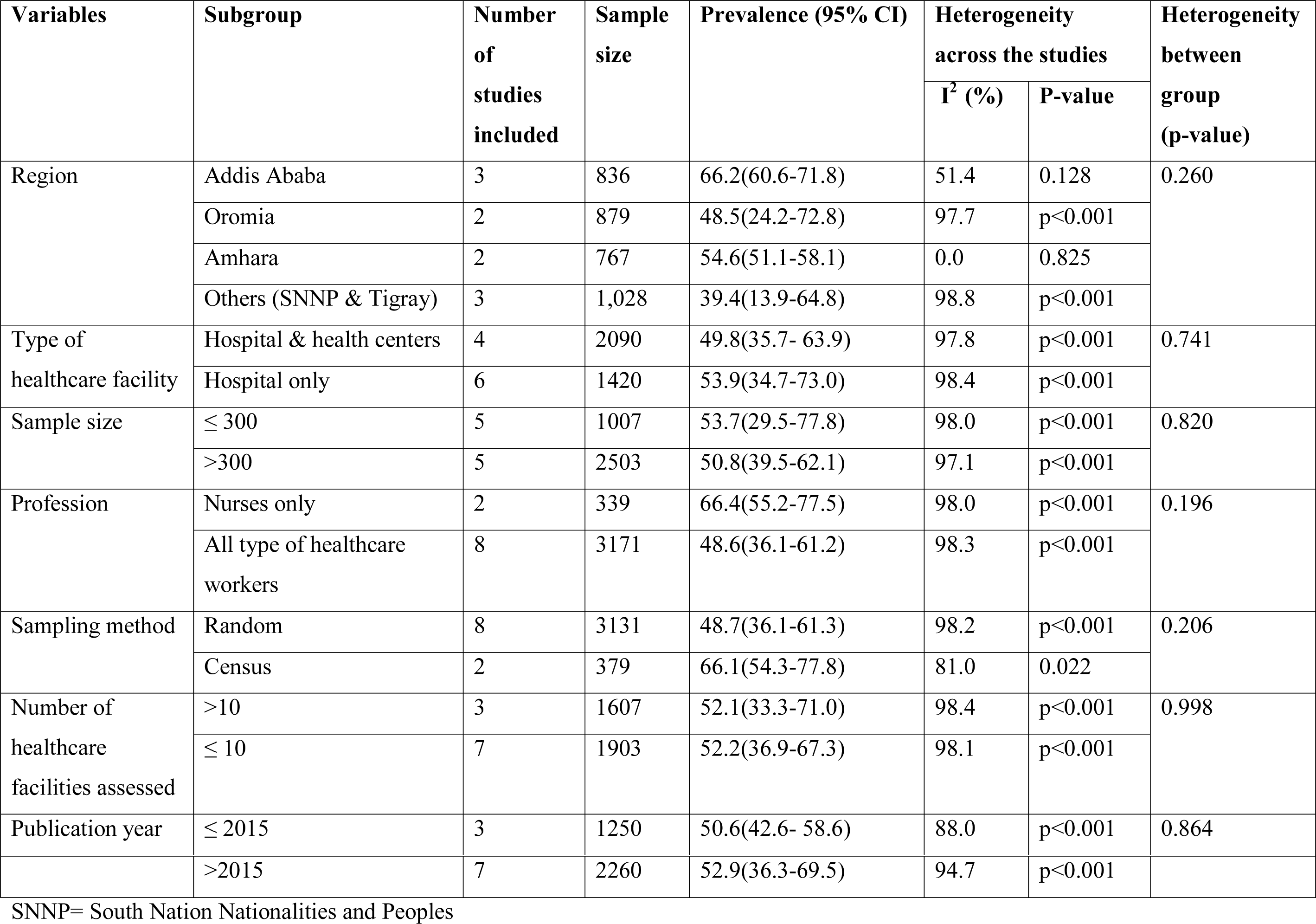
The subgroup prevalence of safe infection prevention practice in Ethiopia, 2014-2019.

### Publication Bias

In the present study, Begg’s and Egger's tests were utilized to detect the presence of publication bias. However, none of the tests revealed significant publication bias (p-values of 0.210 and 0.246, respectively) for the prevalence of safe infection prevention practice in Ethiopia (**Fig 3**).

**Fig 3:**
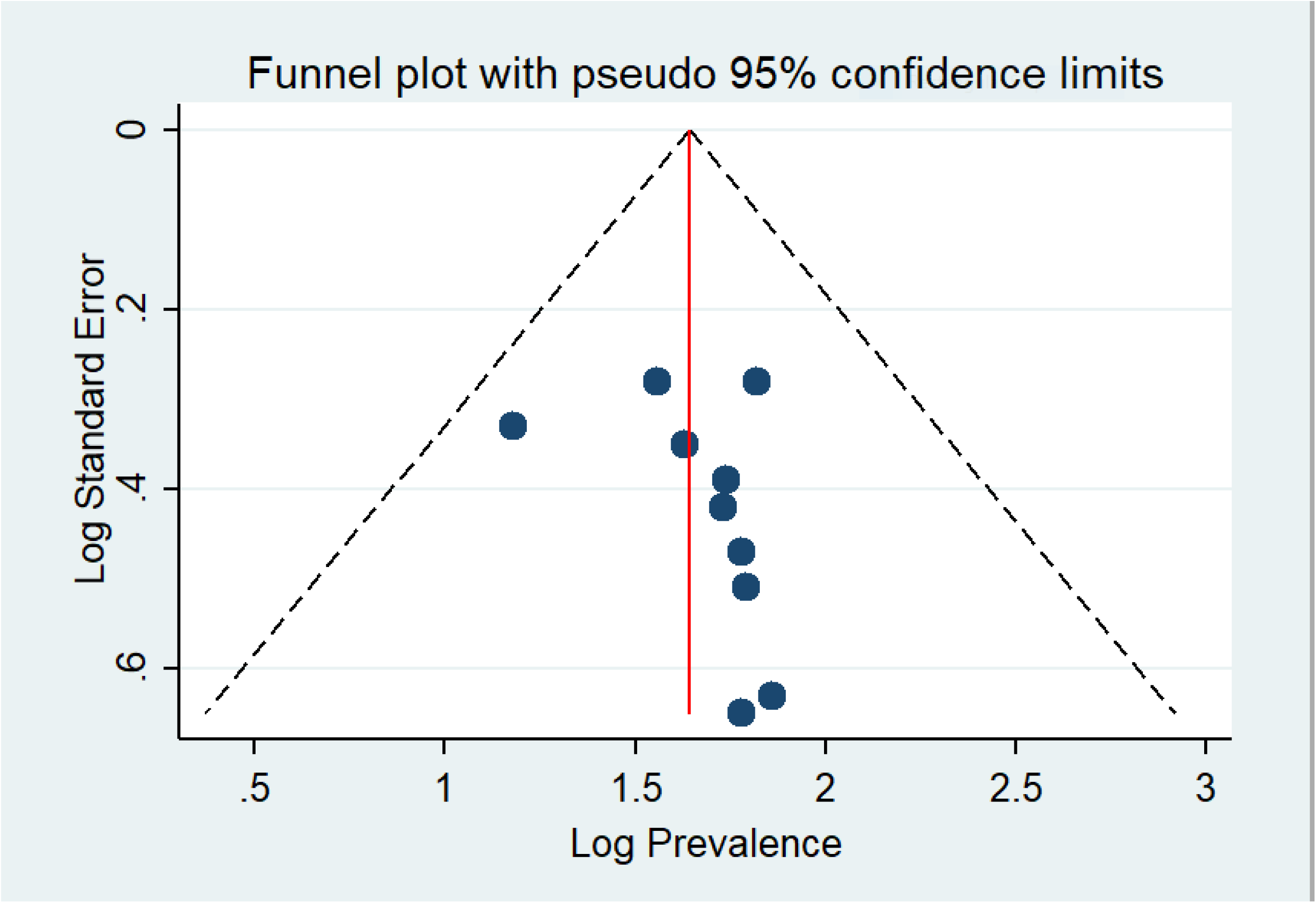
Funnel plot showing publication bias on prevalence studies among healthcare workers in Ethiopian, a systematic review and meta-analysis, Ethiopia

### Sensitivity Analysis

**Table 2** shows the sensitivity analysis of prevalence for each study being removed at a time. To identify the potential source of heterogeneity in the analysis, a leave-one-out sensitivity analysis on the prevalence of infection prevention practice in Ethiopia was employed. The results of this sensitivity analysis showed that the findings were robust and not dependent on a single study. The pooled estimated prevalence of infection prevention practice varied between 56.2 (95%CI: 48.1-64.4) and 50.0 (95%CI: 38.3-61.6) after removing a single study.

Moreover, to identify the possible sources of variations across studies, the meta-regression model was performed by considering the geographical region, publication year, and sample size as covariates. The geographical region (p-value= 0.260), publication year (p-value= 0.864), and sample size (p-value= 0.820) were not statistically significant source of heterogeneity **(Table 3)**.

### Narrative Review

From the ten studies, we summarized descriptively the factors that were associated with safe infection prevention practices in Ethiopia. Factors were categorized into the following three domains: socio-demographic factors (four factors), behavior-related factors (three factors), and healthcare facility-related factors (five factors). The overview of these factors including the strength of association and corresponding articles was presented in **Table 4**.

### Socio-demographic Factors

Four socio-demographic factors were significantly associated with safe infection prevention practices. Healthcare workers age [19,41], gender [33,38], profession [19,20,27,42], and higher service year [31] were identified as underlying factors associated with safe infection prevention practice. The odds of safe infection prevention practices were higher among the age groups between 20-29 [19], 30-39 [19], and 31-40 [41] than HCWs of greater age. The odds of safe infection prevention practices were also higher in female HCWs than males [33,38]. Lastly, significantly lower odds on safe infection prevention practices were observed among all professionals such as midwives [20], laboratory technicians [27], health officers and health assistants [42], and physicians and nurses [19] (**Table 4)**.

### Behavioral related factors

Having good knowledge of infection prevention measures was identified as a factor associated with safe infection prevention practices [27]. In the same way, having a positive attitude towards infection prevention measures, and awareness on infection prevention guideline were the most commonly identified factors associated with the aforementioned practice [27,33] (**Table 4**).

### Healthcare facility related factors

As illustrated in **Table 4**, four healthcare facility-related factors were positively and significantly associated with safe infection prevention practices in Ethiopia. Healthcare workers who worked in facilities with continuous water supply have higher odds on safe infection prevention practice [27]. Similarly, healthcare workers who worked in facilities with access to infection prevention guidelines in the working department have higher odds on the prevention practice [19,20,27]. Lastly, factors such as the type of healthcare facility, current working department, and completion of formal infection prevention training, were the most important factors associated with this prevention practice [20,27,33,38,42].

**Table 4:**
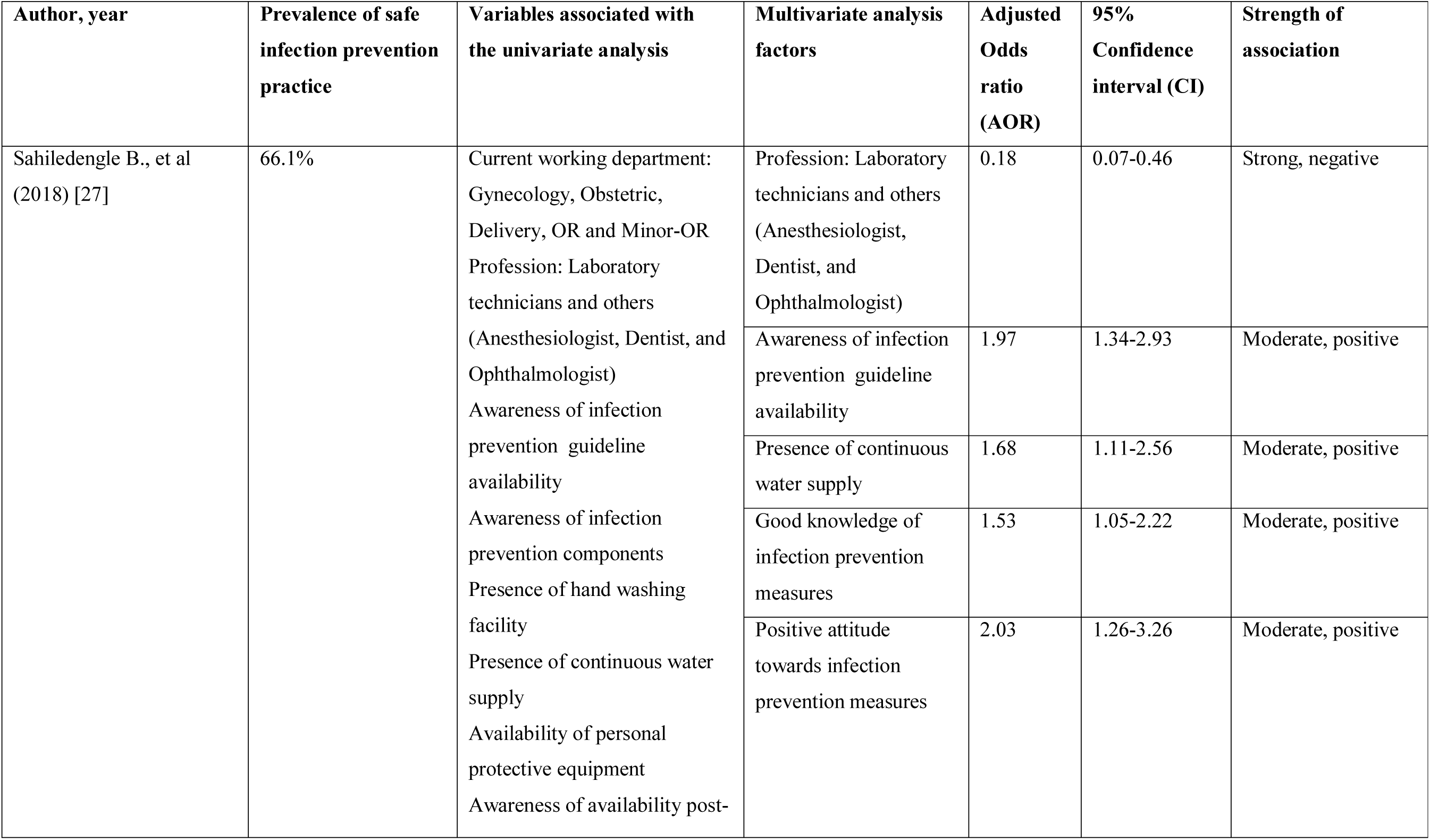
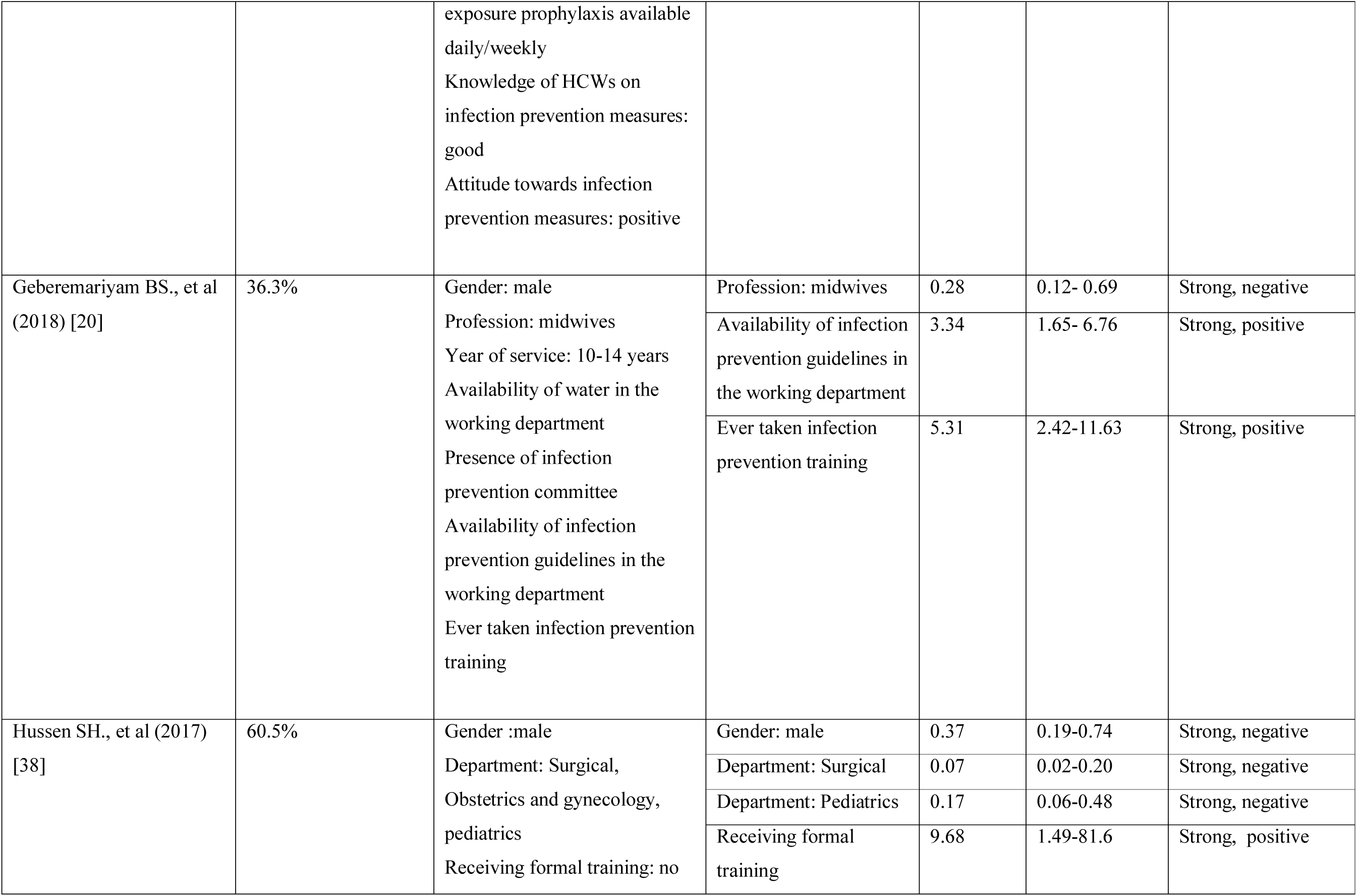
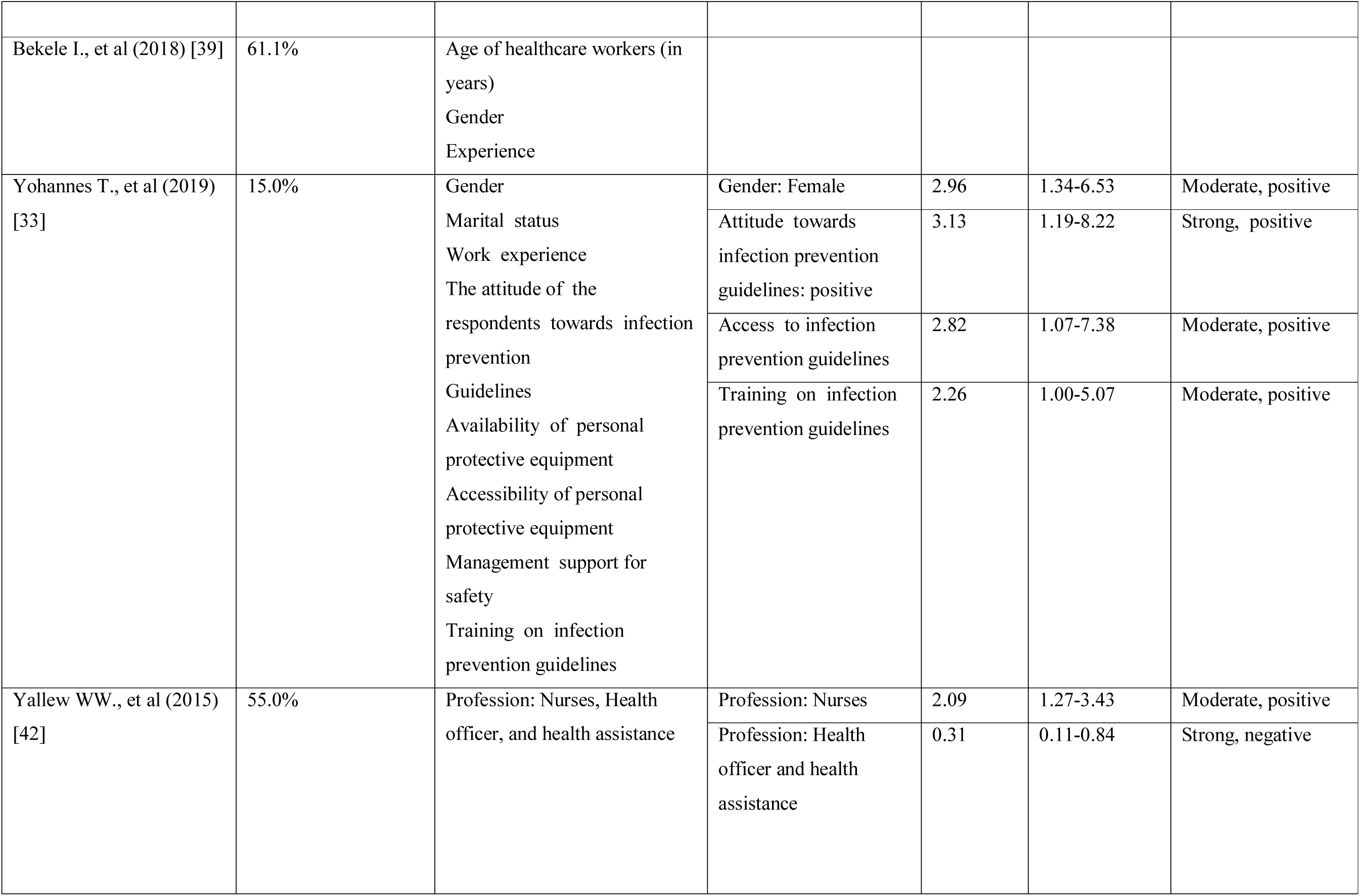
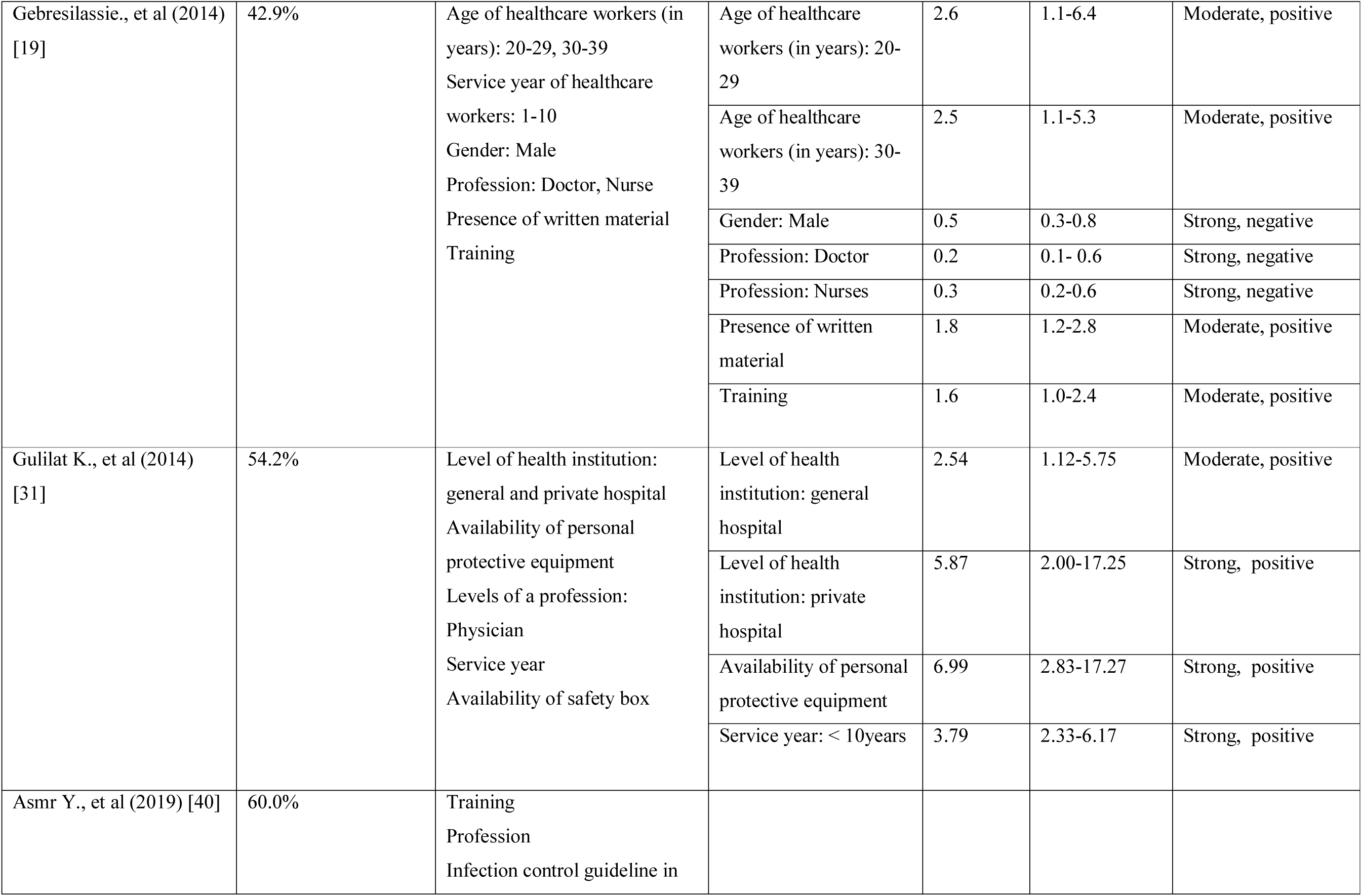
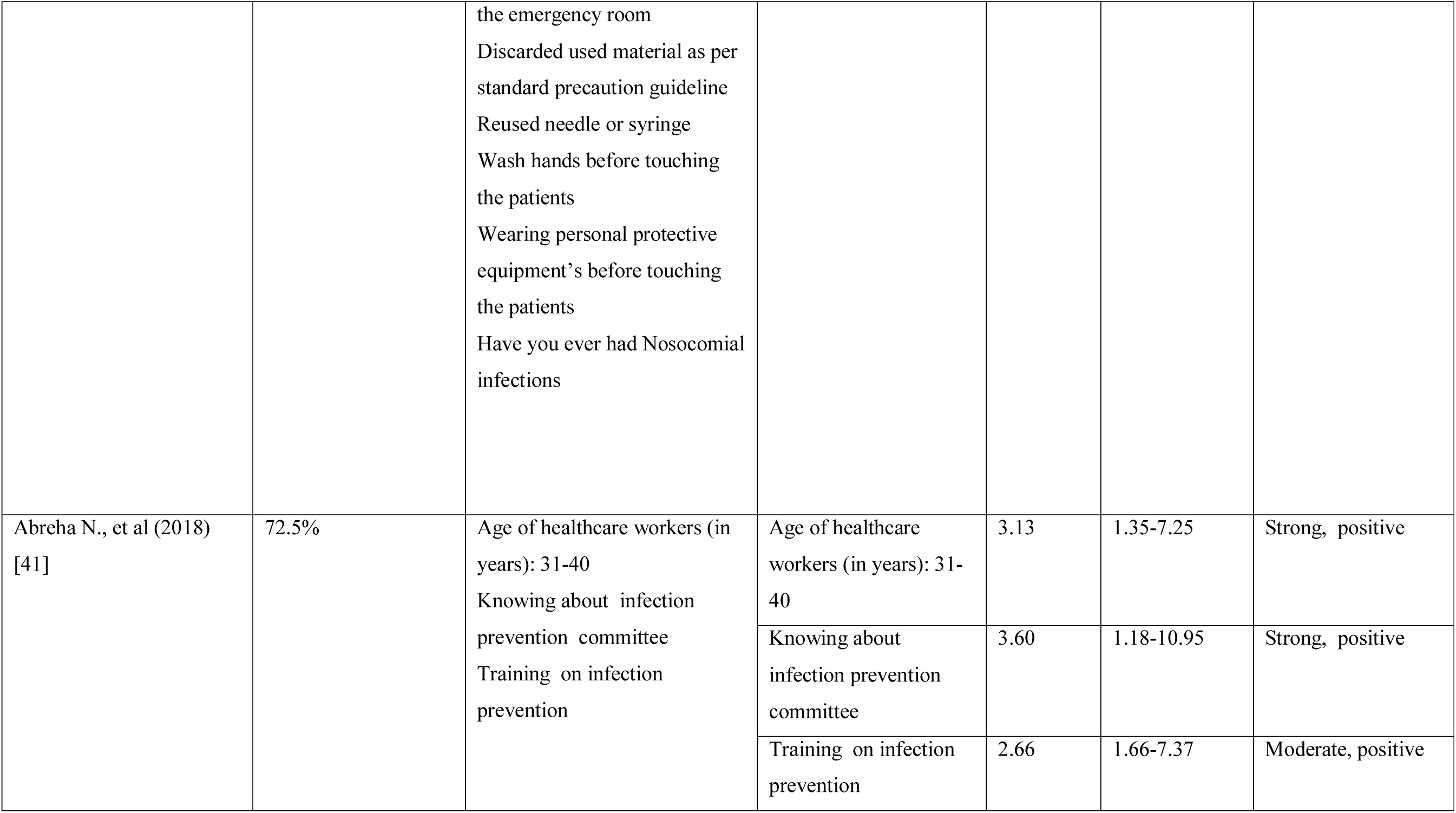
Summary of factors associated with healthcare workers safe infection prevention practice of studies included in Ethiopia, systematic review, 2014-2019.

## Discussion

Infection prevention is a fundamental measure for the control of HAIs. However, sustained compliance from the recommended infection prevention principles among HCWs in developing countries is poor. In Ethiopia, findings regarding the prevalence of safe infection prevention practices have been highly variable. We conducted this systematic review and meta-analysis to estimate the pooled prevalence of safe infection prevention practices among HCWs in Ethiopia. Based on the meta-analysis result, only one-half of the HCWs in Ethiopia had safe infection prevention practices. In our qualitative syntheses, healthcare workers’ socio-demographic, behavioral and healthcare facility-related factors were important variables associated with infection prevention practice.

The result of the 10 included studies noted that the pooled prevalence of safe infection prevention practice in Ethiopia was 52.2% (95%CI: 40.9-63.4). This finding brought important information, and these signified that unsafe practices in healthcare facilities are a major public health concern in Ethiopia. As the burdens of HAIs are increasing [14-18], the current suboptimal infection prevention practices have serious implications to both the HCWs and patients.

On one hand, contracting an infection while in the healthcare facility due to poor infection prevention practice violates the basic idea that healthcare is meant to make people well. In fact, the risk of contracting HAIs is variable and multifaceted: prevalently, it depends on a patient's immune status, the local prevalence of various pathogens, and the HCWs infection prevention practices. Hence, the need for having strong infection prevention programs nationally; and at the healthcare facility levels have been therefore important [29,30,32,43,44]. Un sustained compliance with infection prevention possibly places HCWs at equal, if not at higher risk of contracting bacterial and viral infections such as HIV, HBV, HCV, and MRSA in healthcare facilities [9]. In light of this, studies conducted in Ethiopia even showed a positive correlation between poor standard precaution practices and high prevalence of blood and body fluid exposure [20,27,45,46]. For this reason, the Federal Ministry of Health infection control professionals, healthcare facility administrators, and hospital epidemiologists must pay considerable attention to curve the current poor suboptimal infection prevention practices [47, 48].

In the subgroup analysis, a variation in HCWs infection prevention practices across geographical regions was found. Safe infection prevention practices were consistently more frequent in central Ethiopia (Addis Ababa) and less in Tigray and SNNP regions-the reason for these regional differences may be explained by studies conducted in central Ethiopia included mainly in tertiary and referral hospitals which and commonly staffed are with skilled and experienced healthcare professionals as compared to those in other regions. Another possible explanation for this variation might be due to the difference in environmental infrastructures and behavioral characteristics of HCWs. Our findings may, therefore, indicate the need not only to promote infection prevention and patient safety protocols for the existing HCWs in Ethiopia, moreover to address regional variations through systematic intervention measures.

Our meta-analysis also found that the prevalence of safe infection prevention practices differed between professions-nurses [65.4%, 95%CI (54.5-76.2)] and other healthcare workers [48.4%, 95%CI (35.7-61.0)]. The possible explanation for this observed discrepancy may be due to the training and job description of healthcare workers; the nurses were engaged in inpatient care, and they may have better understanding regarding infection prevention. Still, this prevalence is suboptimal and great concern, therefore, is necessary to strive for a better quality of healthcare.

In this review, the descriptive narration of the included studies on factors associated with safe infection prevention practice was done. Notably, three main domains of determinant factors were identified- socio-demographic, behavioral, and healthcare facility-related factors. From the result, HCWs working in facilities with access to infection prevention guidelines and those receiving formal infection prevention training have higher odds on safe practice. This may, as HCWs infection prevention knowledge, might increase compliance [27]. In this sense, the current systematic review suggests that it may be more effective to improve HCWs infection prevention practice through regular in-service training [49]. A holistic approach is needed to address the scarcity of personal protective equipment and the availability of running water supplies since the availability of these factors also correlates positively with safe infection prevention practices [27,31].

### Limitations of the study

This systematic review and meta-analysis have several limitations. The first limitation considered to conduct this review was to include English language articles only. Second, all of the studies included in this review were cross-sectional as a result; the outcome variable might be affected by other confounding variables. Third, this meta-analysis represented only studies that were reported from the four regions of the country- this irregular distribution of studies around the country may be under-represented and could have limited the study found. Fourth, the majority of the studies included in this review had relatively small sample sizes which could have affected the estimated safe infection prevention practice reports. Fifth, a small number of studies were included in subgroup analyses which reduce the precision of the estimate and a considerable heterogeneity was identified among the studies. Lastly, almost all studies included in this meta-analysis were often based on self-reported data from healthcare providers, which tended to have overestimated compliance and limited the strengths of the findings.

## Conclusions

Infection prevention practices in Ethiopia were significantly low, and only half of the healthcare workers reported safe practices. There were regional and professional variations in the prevalence on the safe practices-it is therefore important for all HCWs to adhere to the existing infection control guidelines strictly, and embedding them in everyday practice. It is also imperative for healthcare administrators to ensure the implementation of infection prevention and patient safety programs in all healthcare settings. Moreover, our study highlights the need to step-up efforts to intensify the current national infection prevention and patient safety initiatives as key policy direction by the Ethiopian Federal Ministry of Health (FMoH). Finally, to be successful, interventions need to be sustained over time in the context of patient safety culture.

## Data Availability

All relevant data are within the manuscript and its Supporting Information files.

## Acknowledgments

The authors would like to thank Madda Walabu University Goba Referral Hospital Public Health Department staff for providing their unreserved support. We would like to thank for the valuable support we received from Mr. John Edward Quisido (assistant professor) as well as Dr. David Allison for their editorial and proofreading support.

## Supporting information

S1 File. Table. PRISMA Checklist

S2 File. Search Strategy (Full searching strategies for PubMed)

S3 File. Search Strategy (Example Google scholar)

S4 File. Table: Methodology quality assessment of included and excluded studies

